# Weight gain during tuberculosis treatment increases the risk of post-tuberculosis metabolic syndrome

**DOI:** 10.1101/2025.11.24.25340881

**Authors:** Argita D. Salindri, Teona Avaliani, Mariam Gujabidze, Leila Goginashvili, Sergo Vashakidze, Zaza Avaliani, Sara C. Auld, Jeffrey M. Collins, Jason R. Andrews, Russell R. Kempker, Hardy Kornfeld, Maia Kipiani, Matthew J. Magee

**Affiliations:** Department of Population Health Sciences, Georgia State University School of Public Health, Hubert Department of Global Health, Emory University Rollins School of Public Health, Atlanta, GA, USA; Division of Infectious Diseases and Geographic Medicine, Department of Medicine, Stanford University School of Medicine, Stanford, CA, USA; National Center for Tuberculosis and Lung Diseases, Tbilisi, Georgia; The University of Georgia, Tbilisi, Georgia; European University, Tbilisi, Georgia; Division of Pulmonary, Allergy, Critical Care, and Sleep Medicine, Department of Medicine, Emory University School of Medicine, Atlanta, Georgia, USA; Department of Epidemiology, Emory University Rollins School of Public Health, Atlanta, Georgia, USA; Division of Infectious Diseases, Department of Medicine, Emory University School of Medicine, Atlanta, GA, USA; Division of Pulmonary, Allergy, and Critical Care, Department of Medicine, University of Massachusetts Medical School, MA, USA; David Tvildiani Medical University, Tbilisi, Georgia; Hubert Department of Global Health, Rollins School of Public Health, Emory University, Atlanta, GA, USA

**Keywords:** tuberculosis, post-TB health, metabolic disease, visceral adipose index

## Abstract

**Background:** Little is known about the relationship between weight gain during treatment and the subsequent risk of cardiovascular and metabolic (cardiometabolic) diseases. We assessed the relationship between changes in body mass index (BMI) during TB treatment with prevalence post-TB metabolic syndrome, a strong predictor of cardiometabolic diseases.

**Methods:** We enrolled a prospective cohort of individuals successfully treated for TB disease in Tbilisi, Georgia, from 2019 – 2022. Eligible participants were HIV-negative individuals aged ≥16 years with newly diagnosed and laboratory-confirmed pulmonary TB. Our study exposure was the relative change in BMI from treatment initiation to treatment completion, dichotomized using ≥5% relative increase cut-off. Our primary study outcome was prevalence post-TB metabolic syndrome (i.e., having ≥3 of the following: elevated blood pressure, elevated triglycerides, low high-density lipoprotein, elevated glycated hemoglobin [HbA1c], and abdominal obesity) at any study visits (including the end of, 6- and 12-months post-TB treatment). Multilevel models were used to estimate the effect of BMI change on post-TB metabolic syndrome.

**Results:** Among 120 participants, the adjusted risk of having metabolic syndrome after TB treatment among those with ≥5% relative increase in BMI was 2.07 times (95% confidence interval [CI] 1.07–4.01) the risk of those with <5% relative increase in BMI during treatment. Additionally, the adjusted mean of post-TB HbA1c among those with ≥5% relative increase in BMI was 0.37 (95%CI 0.03–0.71) points higher compared to those with <5% relative increase in BMI.

**Conclusions:** Our findings indicate that weight gain during TB treatment may influence the risk of cardiovascular/metabolic diseases after TB treatment.

**Key points summary:** In a cohort of persons successfully treated for pulmonary tuberculosis, nearly half had ≥5% BMI increase during treatment. Notably, those with ≥5% BMI increase had higher glycated hemoglobin levels and were twice as likely to have metabolic syndrome after tuberculosis treatment completion.

## INTRODUCTION

Post-TB sequelae, defined as pathological conditions following TB treatment, may impact a majority of the ∼155 million prevalent TB survivors. [1, 2] Mounting evidence suggests that TB survivors experience residual complications and have high post-TB mortality rates. [3] Importantly, an estimated 20% of post-TB mortality is attributable to cardiovascular diseases. [3, 4] Despite increased attention to post-TB health, the field lacks key clinical data (e.g., TB-associated factors), and there are no established guidelines to prevent cardiovascular health outcomes among TB survivors. Furthermore, there is a general lack of knowledge about risk factors for adverse cardiovascular and metabolic (cardiometabolic) health post-TB. Identification of factors associated with increased post-TB cardiometabolic diseases would help guide preventive efforts to reduce mortality rates and prevent chronic sequelae among TB survivors.

While weight gain during TB treatment is generally beneficial, [5] it is unclear to what extent weight gain during TB treatment impacts post-TB cardiometabolic health. Low body mass index (i.e., BMI <18.5kg/m^2^) is a well-established risk factor for TB disease, and a common syndrome associated with TB is cachexia, resulting in low BMI at TB diagnosis. [6–8] Weight gain during treatment is associated with improved TB treatment outcomes, including fewer adverse events and lower mortality. [5, 9, 10] However, the rapid weight gain that may be experienced by some individuals during TB treatment is associated with adverse cardiovascular outcomes in other contexts. For instance, a rapid weight gain (i.e., ∼4.5 kg increase in body weight over a year) among individuals with type-2 diabetes mellitus (diabetes) or stroke is associated with increased risks of cardiovascular events, including death. [11, 12] Whether this established relationship between weight gain and cardiovascular outcomes is observed among individuals with TB is unknown.

Metabolic syndrome is a cluster of clinical markers of metabolic dysfunction associated with cardiometabolic diseases, including diabetes, stroke, and heart disease. However, most studies of post-TB health are limited to retrospective study designs and do not characterize changes in cardiometabolic disease risks. Characterizing the association between weight gain during TB treatment and post-TB metabolic syndrome may help estimate the risk of cardiometabolic disease progression among nearly 9 million new TB survivors annually. Thus, we conducted a cohort study among participants with pulmonary drug-susceptible and drug-resistant TB to describe changes in cardiometabolic markers following successful TB treatment. We also determined if the relative change of BMI during TB treatment was associated with post-TB metabolic syndrome.

## METHODS

### Study Design and Setting

We conducted a 12-month prospective cohort study among individuals with drug-susceptible and drug-resistant TB who completed treatment and received “microbial cure” status at the end of treatment in Tbilisi, Georgia, from December 2019 – July 2022. Participants ≥16 years old with newly diagnosed, bacteriologically confirmed pulmonary TB and available information on BMI change during treatment (i.e., retrospectively collected by reviewing participants’ medical charts) were eligible for study inclusion. Individuals with a history of lung surgery or lung cancer before the current TB episode, relapse or retreatment cases, pregnant women, and people living with HIV co-infection were excluded from the final analyses. We also excluded individuals whose treatment duration was >8 months (for drug-sensitive or isoniazid-monoresistant TB [DS/HR TB]) or >24 months (for rifampicin- or multidrug/extensively drug-resistant TB [RR or M/XDR TB]).

We enrolled individuals who completed TB treatment and had favorable outcomes (i.e., completed treatment or received microbial cure status [13]) at the end of treatment and conducted study visits with participants at the end of treatment, 6 months, and 12 months after TB treatment completion. Anthropometric measurements (e.g., body weight, height, blood pressure, waist circumference), lipid profiles (i.e., high-density lipoprotein [HDL], low-density lipoprotein [LDL], total cholesterol, triglycerides), and glycated hemoglobin (HbA1c) were measured at each visit (i.e., end of TB treatment, 6- and 12-month follow-ups). A study questionnaire was administered at each visit. All blood assays were analyzed at the NCTLD laboratory facility using BioSystem A25 Analyzer (Barcelona, Spain).

### Definitions and Study Measures

Our primary exposure was relative change in BMI during TB treatment, expressed in a percentage that represents the absolute change in BMI (i.e., obtained by subtracting BMI values measured at the end of TB treatment from values measured at TB treatment initiation) relative to values at TB treatment initiation. We then used a 5% cut-off to classify the relative change in BMI into “low” (i.e., relative BMI change <5%) or “high” (i.e., relative BMI change ³5%). We selected the 5% cut-off because the median relative change in BMI in our cohort was 4.7% and two previous studies have used 5% weight gain threshold, including one that used it to predict TB mortality. [14, 15] In our study, individuals with reduced BMI (n = 15) were grouped into the “low BMI change” category due to the small sample size.

The primary study outcome was metabolic syndrome independently measured at the end of TB treatment, 6 months, and 12 months post-TB treatment visits. We defined metabolic syndrome as having ≥3 of the following conditions: 1) elevated blood pressure (a single reading of systolic blood pressure [SBP] ≥130 mmHg and/or diastolic blood pressure [DBP] ≥80 mmHg), 2) elevated triglycerides (triglyceride ≥150 mg/dL), 3) low HDL (i.e., HDL <40 mg/dL for male and <50 mg/dL for female), 4) elevated blood glucose (HbA1c ≥5.7%), and 5) abdominal obesity, defined as waist circumference of ≥88 cm in women and ≥102 cm in men; following the American Heart Association guidelines. [16] We also assessed the association between change in BMI and the individual components of metabolic syndrome, as well as total cholesterol, LDL, and visceral adipose index (VAI). VAI is a measure of visceral fat associated with cardiometabolic risks and was calculated as previously described. [17–19]

Other key variables included demographic characteristics and behavioral risk factors (i.e., smoking, alcohol, and drug use) were collected at the time of study enrollment and follow-up visits. Clinical information at the beginning of- or during TB treatment (e.g., drug-resistance profile, BMI at TB treatment initiation, time to achieve sputum conversion, lung abnormalities, and comorbidity factors) were abstracted from medical charts. For our study, individuals with a) drug-susceptible TB [DSTB] and b) *Mycobacterium tuberculosis (Mtb)* strain resistant to isoniazid and susceptible to rifampicin (HR TB) were grouped as “DS/HR TB” as the majority of those with HR TB had low-level isoniazid resistance and received treatment similar to those with DSTB; while patients with *Mtb* strains resistant to at least rifampicin were grouped as “RR or M/XDR TB” (i.e., included individuals with rifampicin resistant-, multidrug resistant-, and extensively drug resistant-TB). Study data were collected and managed using Research Electronic Data Capture (REDCap, Vanderbilt University, NC) electronic data capture tools hosted at Emory University. [20, 21]

### Statistical Analyses

We performed Chi-square/Fisher’s exact test and Wilcoxon rank-sum test to assess the association between participant characteristics and change in BMI status. We used modified Poisson with generalized estimating equations (MP-GEE), a marginal modeling framework for non-linear measures that accounts for within-subject correlations due to the repeated measures, to estimate the unadjusted and adjusted risk ratios (with corresponding 95%CI) of dichotomous study outcomes: metabolic syndrome, elevated blood pressure levels, elevated triglycerides, low HDL, elevated blood glucose, or abdominal obesity at the end of TB treatment and during post-TB visits. Similarly, we performed general linear mixed models with random intercept to estimate the association between change in BMI during treatment and post-TB levels of cardiometabolic markers (i.e., HbA1c, LDL, HDL, total cholesterol, triglycerides, and VAI). We also evaluated whether the association between increases in BMI during TB treatment and post-TB metabolic syndrome was modified by baseline BMI and diabetes status. Covariates included in the multivariable models were identified by assessing bivariate associations, directed acyclic graph theory[22], and previously published literature. All analyses were performed in SAS version 9.4 (Cary, North Carolina) with p-values <0.05 considered statistically significant. Figures were generated in R (version 4.3.3) using *“ggplot2”* package. [23, 24]

### Sensitivity analyses

A sensitivity analysis was performed to quantify systematic errors due to misclassification of metabolic syndrome. To account for misclassifications of metabolic syndrome, we performed additional analyses where we accounted for self-reported use of prescription medication to manage metabolic diseases (i.e., recorded in the medical chart or reported during the study questionnaire administration) to define relevant components of metabolic syndrome. For example, we defined lower HDL as either having an HDL level <40 mg/dL for males and <50 mg/dL for females or currently taking statin or other medications to lower blood lipid levels. [25] We also performed multiple GEE and linear mixed models to account for potential systematic errors due to covariate misspecifications in the multivariable models.

## RESULTS

During the study period, we enrolled a total of 140 patients who were successfully treated for TB disease, 20 of whom were excluded due to missing BMI values at the beginning of TB treatment or other reasons **(Figure S1)**; leaving 120 (85.7%) individuals included in the final analyses. By the end of the study period, 115 participants remained in the cohort (loss to follow-up rate of 4.2%). Among included participants, the majority were male (59.2%), Georgian (90.0%), and the median age at study enrollment was 37 (interquartile range [IQR] 27 – 49). The prevalence of RR or M/XDR TB was 23.3% and cavitary disease at treatment initiation was identified in 22.5% of study participants **(Table 1)**.

**Table 1.**
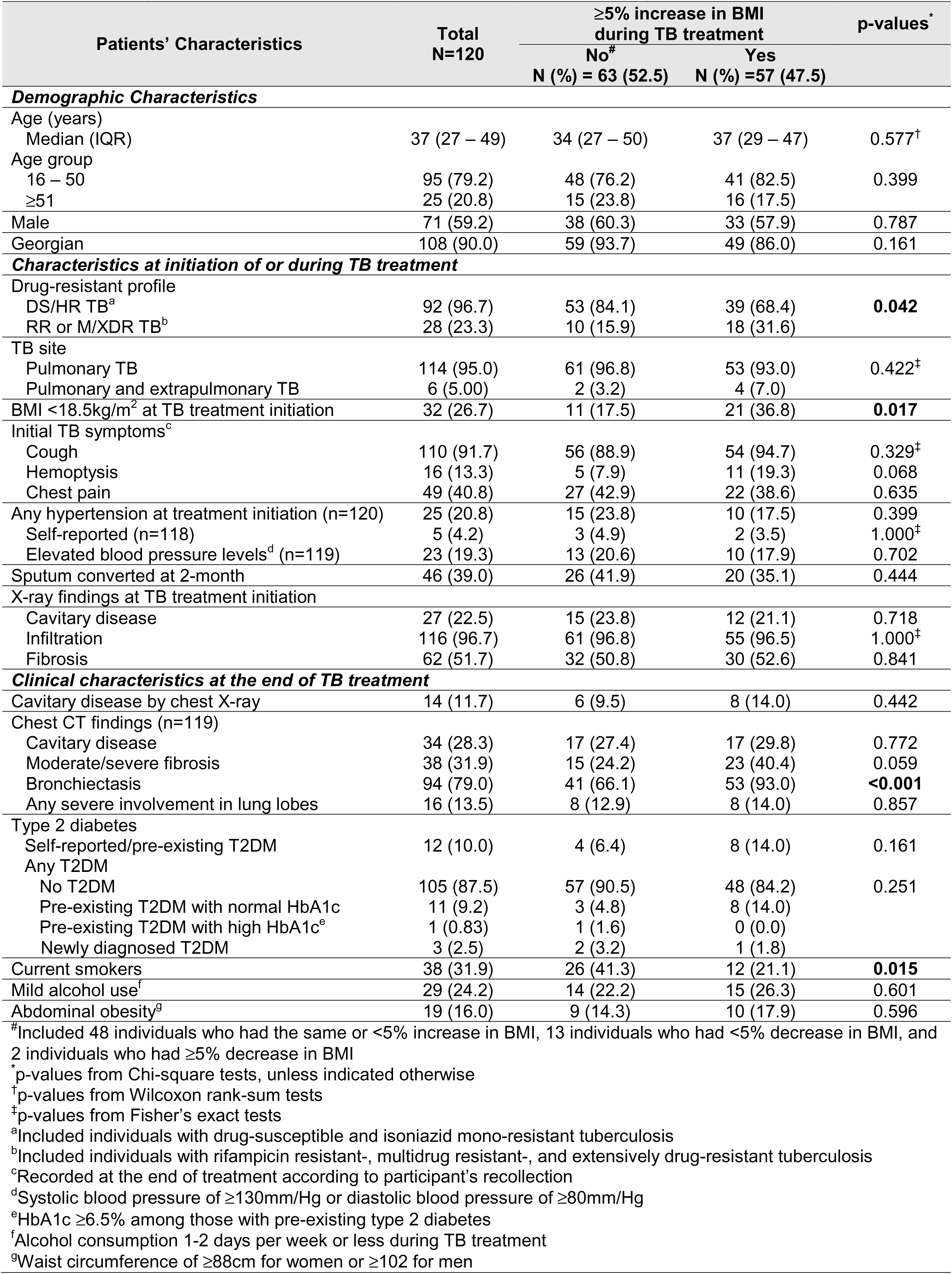
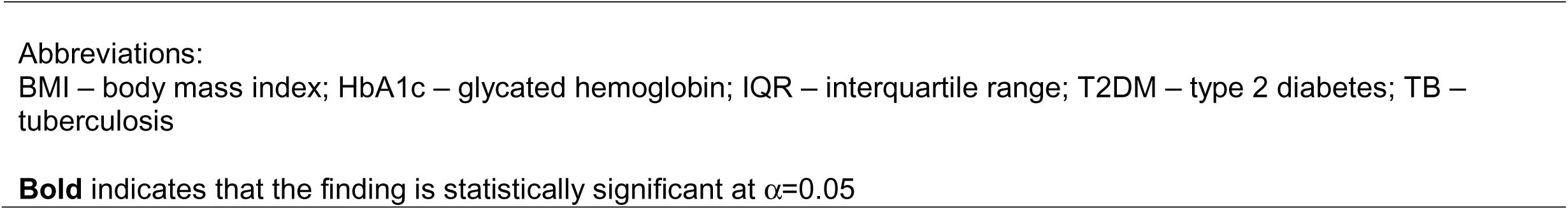
Demographic and clinical characteristics of individuals who were successfully treated for pulmonary tuberculosis disease according to changes in body mass index during treatment in the country of Georgia, 2019 – 2022 (N=120)

| Patients' Characteristics | Total<br>N=120 | ≥5% increase in BMI<br>during TB treatment |  | p-values* |
| --- | --- | --- | --- | --- |
|  |  | No<sup>#</sup><br>N (%) = 63 (52.5) | Yes<br>N (%) =57 (47.5) |  |
| **Demographic Characteristics** |  |  |  |  |
| Age (years) |  |  |  |  |
| Median (IQR) | 37 (27 – 49) | 34 (27 – 50) | 37 (29 – 47) | 0.577<sup>†</sup> |
| Age group |  |  |  |  |
| 16 – 50 | 95 (79.2) | 48 (76.2) | 41 (82.5) | 0.399 |
| ≥51 | 25 (20.8) | 15 (23.8) | 16 (17.5) |  |
| Male | 71 (59.2) | 38 (60.3) | 33 (57.9) | 0.787 |
| Georgian | 108 (90.0) | 59 (93.7) | 49 (86.0) | 0.161 |
| **Characteristics at initiation of or during TB treatment** |  |  |  |  |
| Drug-resistant profile |  |  |  |  |
| DS/HR TB<sup>a</sup> | 92 (96.7) | 53 (84.1) | 39 (68.4) | **0.042** |
| RR or M/XDR TB<sup>b</sup> | 28 (23.3) | 10 (15.9) | 18 (31.6) |  |
| TB site |  |  |  |  |
| Pulmonary TB | 114 (95.0) | 61 (96.8) | 53 (93.0) | 0.422<sup>‡</sup> |
| Pulmonary and extrapulmonary TB | 6 (5.00) | 2 (3.2) | 4 (7.0) |  |
| BMI <18.5kg/m<sup>2</sup> at TB treatment initiation | 32 (26.7) | 11 (17.5) | 21 (36.8) | **0.017** |
| Initial TB symptoms<sup>c</sup> |  |  |  |  |
| Cough | 110 (91.7) | 56 (88.9) | 54 (94.7) | 0.329<sup>‡</sup> |
| Hemoptysis | 16 (13.3) | 5 (7.9) | 11 (19.3) | 0.068 |
| Chest pain | 49 (40.8) | 27 (42.9) | 22 (38.6) | 0.635 |
| Any hypertension at treatment initiation (n=120) | 25 (20.8) | 15 (23.8) | 10 (17.5) | 0.399 |
| Self-reported (n=118) | 5 (4.2) | 3 (4.9) | 2 (3.5) | 1.000<sup>‡</sup> |
| Elevated blood pressure levels<sup>d</sup> (n=119) | 23 (19.3) | 13 (20.6) | 10 (17.9) | 0.702 |
| Sputum converted at 2-month | 46 (39.0) | 26 (41.9) | 20 (35.1) | 0.444 |
| X-ray findings at TB treatment initiation |  |  |  |  |
| Cavitary disease | 27 (22.5) | 15 (23.8) | 12 (21.1) | 0.718 |
| Infiltration | 116 (96.7) | 61 (96.8) | 55 (96.5) | 1.000<sup>‡</sup> |
| Fibrosis | 62 (51.7) | 32 (50.8) | 30 (52.6) | 0.841 |
| **Clinical characteristics at the end of TB treatment** |  |  |  |  |
| Cavitary disease by chest X-ray | 14 (11.7) | 6 (9.5) | 8 (14.0) | 0.442 |
| Chest CT findings (n=119) |  |  |  |  |
| Cavitary disease | 34 (28.3) | 17 (27.4) | 17 (29.8) | 0.772 |
| Moderate/severe fibrosis | 38 (31.9) | 15 (24.2) | 23 (40.4) | 0.059 |
| Bronchiectasis | 94 (79.0) | 41 (66.1) | 53 (93.0) | **<0.001** |
| Any severe involvement in lung lobes | 16 (13.5) | 8 (12.9) | 8 (14.0) | 0.857 |
| Type 2 diabetes |  |  |  |  |
| Self-reported/pre-existing T2DM | 12 (10.0) | 4 (6.4) | 8 (14.0) | 0.161 |
| Any T2DM |  |  |  |  |
| No T2DM | 105 (87.5) | 57 (90.5) | 48 (84.2) | 0.251 |
| Pre-existing T2DM with normal HbA1c | 11 (9.2) | 3 (4.8) | 8 (14.0) |  |
| Pre-existing T2DM with high HbA1c<sup>e</sup> | 1 (0.83) | 1 (1.6) | 0 (0.0) |  |
| Newly diagnosed T2DM | 3 (2.5) | 2 (3.2) | 1 (1.8) |  |
| Current smokers | 38 (31.9) | 26 (41.3) | 12 (21.1) | **0.015** |
| Mild alcohol use<sup>f</sup> | 29 (24.2) | 14 (22.2) | 15 (26.3) | 0.601 |
| Abdominal obesity<sup>g</sup> | 19 (16.0) | 9 (14.3) | 10 (17.9) | 0.596 |

<sup>#</sup>Included 48 individuals who had the same or <5% increase in BMI, 13 individuals who had <5% decrease in BMI, and 2 individuals who had ≥5% decrease in BMI
\*p-values from Chi-square tests, unless indicated otherwise
<sup>†</sup>p-values from Wilcoxon rank-sum tests
<sup>‡</sup>p-values from Fisher's exact tests
<sup>a</sup>Included individuals with drug-susceptible and isoniazid mono-resistant tuberculosis

| Patients' Characteristics | Total<br>N=120 | ≥5% increase in BMI<br>during TB treatment |  | p-values<sup>*</sup> |
| --- | --- | --- | --- | --- |
|  |  | No<sup>#</sup> | Yes |  |
|  |  | N (%) = 63 (52.5) | N (%) =57 (47.5) |  |

Abbreviations: BMI – body mass index; HbA1c – glycated hemoglobin; IQR – interquartile range; T2DM – type 2 diabetes; TB – tuberculosis
**Bold** indicates that the finding is statistically significant at $\alpha$ =0.05

More than a quarter (32/120, 26.7%) of our study participants had a BMI <18.5 kg/m^2^ at TB treatment initiation, while 55.8% (67/120) had BMI 18.5 – 24.9 kg/m^2^, and 17.5% (21/120) had BMI ≥25.0 kg/m^2^ (**Table 1)**. The median relative change in BMI during TB treatment among our cohort was 4.7% (IQR 0 – 10.3). Nearly half (47.5%, 57/120) of study participants gained at least 5% in BMI during TB treatment, while 52.5% (63/120) did not. Of the individuals who did not gain at least 5% in BMI, 48 (76.2%, 48/63) individuals had no change or <5% relative increase in BMI, and 15 (23.8%, 15/63) others had a decreased BMI (two of whom had >5% decrease). Individuals who gained ≥5% relative increase in BMI during TB treatment were more likely to have RR or M/XDR TB and were underweight at TB treatment initiation, but less likely to report current smoking (p<0.05). Individuals’ BMI trajectories from TB treatment initiation to 12-month post-TB treatment are depicted in **Figure S2**.

### BMI change during TB treatment and post-TB metabolic syndrome

Overall, 10.0% of study participants had prevalent metabolic syndrome at the end of TB treatment, 15.8% 6-months post-treatment, and 22.6% 12-months post-treatment **(Table S1)**. The prevalence of metabolic syndrome among those with ≥5% relative increase in BMI was 12.3% at the end of TB treatment, 24.6% at 6-month, and 30.2% at 12-month follow up versus 7.9% at the end of TB treatment, 7.9% at 6-month, and 16.1% at 12-month follow-up among those with <5% relative increase in BMI **(Figure 1)**. The risk of having post-TB metabolic syndrome among those with a high increase in BMI was 2.07 times (95%CI 1.07 – 4.01) the risk of those with a low increase in BMI, after adjusting for age, sex, drug-resistant TB, and the clustering of metabolic syndrome data at the individual level **(Table 2)**.

**Figure 1.**
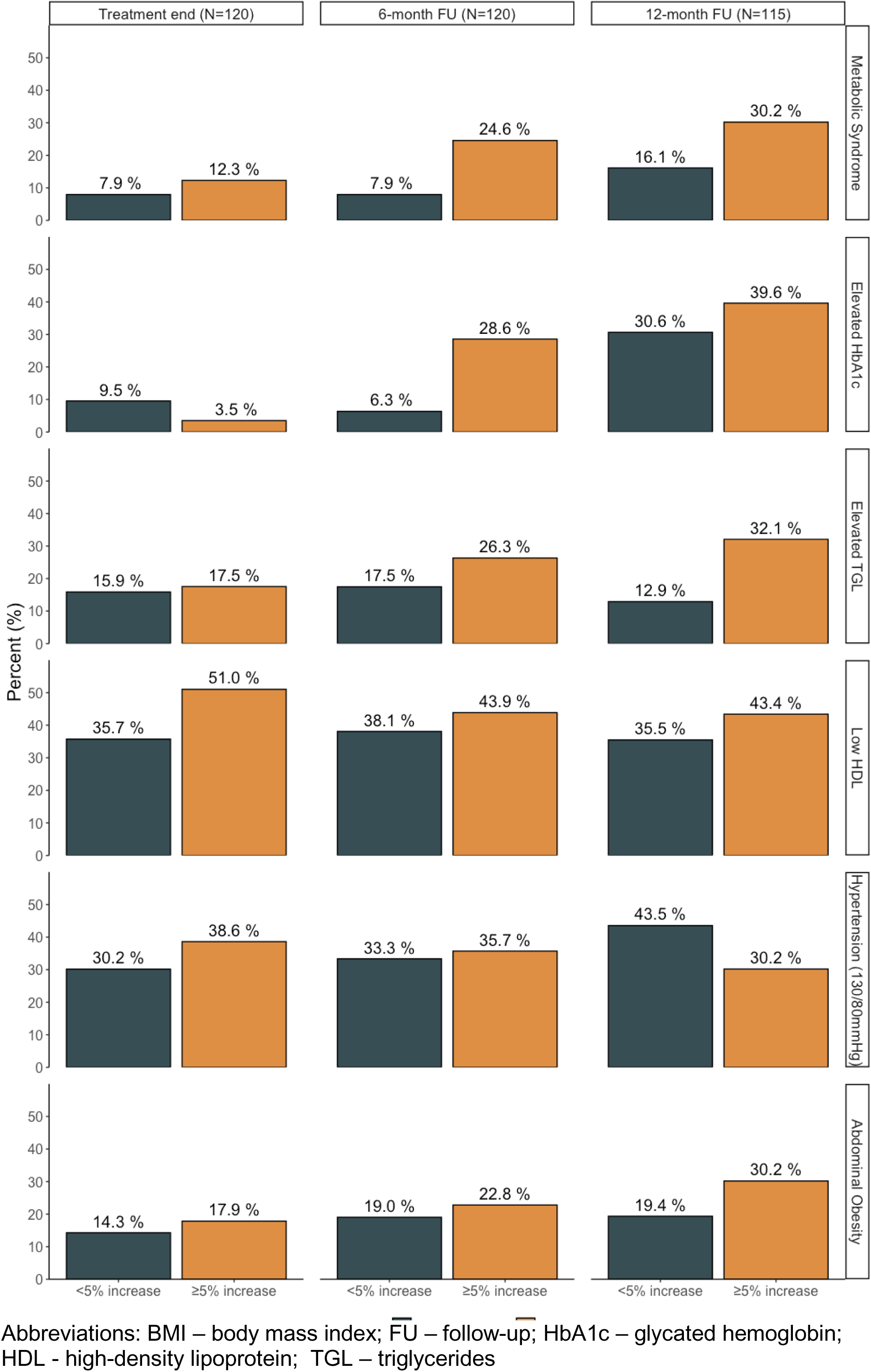
Proportion of post-treatment metabolic syndrome among individuals who were successfully treated for pulmonary tuberculosis disease in the country of Georgia, 2019 – 2022 **Alt text:** Graphs comparing the proportion of post-TB cardiometabolic markers between individuals with ≥5% versus <5% increase in relative BMI at each study visits

**Table 2.** BMI change during treatment and risk of post-treatment metabolic syndrome among individuals who were successfully treated for pulmonary tuberculosis disease in the country of Georgia, 2019 – 2022 (N=120)

| Patients Characteristics | Metabolic syndrome* | Hypertension (≥130/80 mmHg) | Triglycerides ≥150mg/dL | Lower HDL‡ | HbA1c ≥5.7% | Abdominal obesity† |
| --- | --- | --- | --- | --- | --- | --- |
|  | aRR§ (95%CI) | aRR§ (95%CI) | aRR§ (95%CI) | aRR§ (95%CI) | aRR§ (95%CI) | aRR§ (95%CI) |
| BMI Change |  |  |  |  |  |  |
| <5% increase | Reference | Reference | Reference | Reference | Reference | Reference |
| ≥5% increase | **2.07 (1.07 – 4.01)** | 1.12 (0.78 – 1.61) | 1.61 (0.93 – 2.77) | 1.22 (0.92 – 1.62) | 1.50 (0.93 – 2.43) | 1.24 (0.69 – 2.25) |
| Age group |  |  |  |  |  |  |
| 16 – 50 | Reference | Reference | Reference | Reference | Reference | Reference |
| 51+ | **3.22 (1.88 – 5.51)** | **2.79 (2.02 – 3.87)** | 1.45 (0.84 – 2.53) | 1.07 (0.80 – 1.44) | **1.95 (1.23 – 3.13)** | **2.46 (1.47 – 4.11)** |
| Sex |  |  |  |  |  |  |
| Female | Reference | Reference | Reference | Reference | Reference | Reference |
| Male | 0.94 (0.53 – 1.69) | 1.33 (0.91 – 1.94) | 1.74 (0.98 – 4.66) | 1.05 (0.80 – 1.38) | 1.28 (0.79 – 2.07) | **0.45 (0.35– 0.82)** |
| Type of resistance |  |  |  |  |  |  |
| DSTB | Reference | Reference | Reference | Reference | Reference | Reference |
| DRTB | 1.55 (0.86 – 2.80) | 0.94 (0.64 – 1.38) | 1.55 (0.91 – 2.66) | 1.22 (0.90 – 1.66) | **1.73 (1.10 – 2.73)** | 1.31 (0.74 – 2.31) |

\*Having at least 3 of the following conditions: 1) elevated blood pressure, 2) elevated triglycerides, 3) low high-density lipoprotein, 4) elevated blood glucose, and 5) abdominal obesity
†Waist circumference of ≥88cm for women or ≥102 for men
‡HDL <40 mg/dL for male and <50 mg/dL for female
§Results from marginal models using modified Poisson with generalized estimating equations; models were adjusted for age, sex, and type of drug resistance
Abbreviations:
aOR – adjusted risk ratio; BMI – body mass index; CI – confidence interval; DSTB – drug-susceptible tuberculosis; DRTB – drug-resistant tuberculosis
**Bold** indicated that the finding is statistically significant at $\alpha=0.05$

### BMI change and individual components of metabolic syndrome

The prevalence and trajectories of individual components of metabolic syndrome at the end of TB treatment and during post-TB follow-ups are shown in **Tables S1 and S2**. The most common metabolic abnormality observed at the end of TB treatment and during post-TB follow-up visits was low HDL cholesterol, with a prevalence of 42.9% at the end of TB treatment, 40.8% at 6-month post-TB treatment, and 39.1% at 12-month post-TB treatment **(Table S1)**. The mean of HbA1c, HDL, LDL, and total cholesterol levels increased from the end of TB treatment, 6-month, and 12-month post-TB treatment, but the mean of triglyceride and VAI levels were fluctuated, with a slight increase at 6-months post-TB treatment and reverted at 12-months post-TB treatment (e.g., VAI was increasing from 3.61 [standard deviation, SD 2.79] at the end of TB treatment to 4.18 [SD 3.45] at 6-month and 3.73 [SD 2.75] at 12-month post-TB) **(Table S2)**.

The mean HbA1c among those with ≥5% relative increase in BMI was 4.29% (SD 0.90) at the end of TB treatment, 5.33% (SD 1.68) at 6-months, and 5.52% (SD 1.95) at 12-months post-TB versus 4.47% (SD 1.22) at the end of TB treatment, 4.31% (SD 1.21) at 6-months, and 5.21% (SD 1.12) at 12-months post-TB among those with <5% relative increase in BMI **(Figure 2)**. From the multilevel model, the mean HbA1c among those with ≥5% relative increase in BMI was 0.37 percentage points higher (95%CI 0.03 – 0.71) compared to those with <5% relative increase in BMI, after adjusting for age, sex, drug-resistant TB, and the clustering of HbA1c levels at the individual level **(Table 3)**.

**Figure 2.**
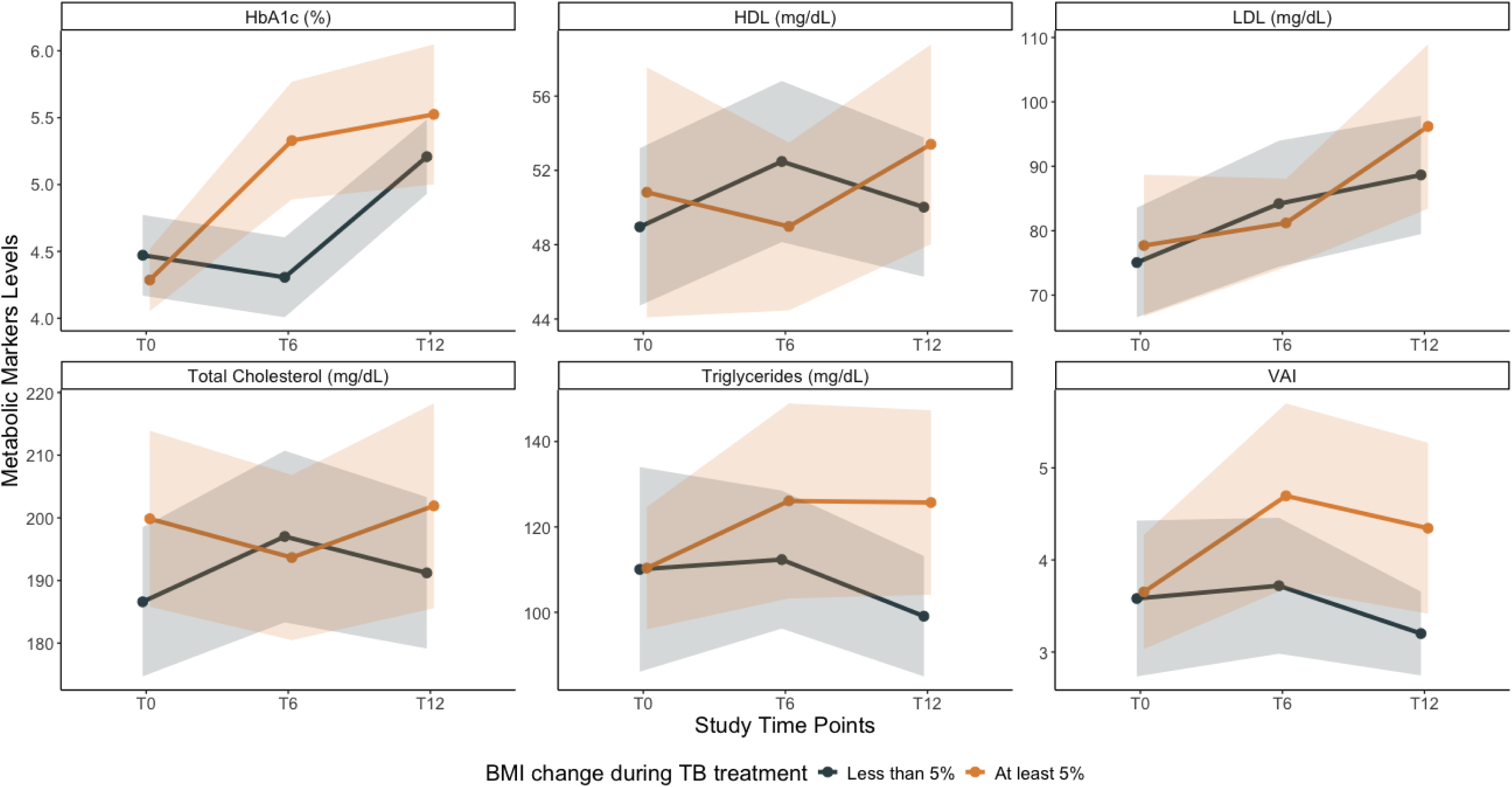
Means of post-treatment metabolic markers among individuals who were successfully treated for pulmonary tuberculosis disease in the country of Georgia, 2019 – 2022 (N=120) **Alt text:** Graphs comparing the trend of post-TB cardiometabolic markers between individuals with ≥5% versus <5% increase in relative BMI at each study visits

**Table 3.** Multilevel analysis for modeling post-treatment levels of glycated hemoglobin (HbA1c), high-density lipoprotein (HDL), low-density lipoprotein (LDL), triglycerides, total cholesterol, and visceral adipose index (VAI), as a function of BMI change during TB treatment, age, sex, type of drug resistance while accounting for repeated measures data among individuals who were successfully treated for pulmonary tuberculosis diseases in the country of Georgia, 2019 – 2022 (N=120)

| Characteristics | Estimates (se) |  |  |  |  |  |
| --- | --- | --- | --- | --- | --- | --- |
|  | HbA1c | HDL | LDL | Total Cholesterol | Triglycerides | VAI |
| Intercept | 4.83 (0.16)*** | 56.03 (2.56)*** | 88.73 (5.36)*** | 190.04 (7.81)*** | 83.45 (11.23)*** | 2.92 (0.45)*** |
| BMI Change |  |  |  |  |  |  |
| <5% increase | Reference | Reference | Reference | Reference | Reference | Reference |
| ≥5% increase | **0.37 (0.17)\*** | 0.49 (2.36) | 2.65 (5.10) | 9.68 (7.67) | 12.51 (10.96) | 0.80 (0.44) |
| Age group |  |  |  |  |  |  |
| 16 – 50 | Reference | Reference | Reference | Reference | Reference | Reference |
| 51+ | **0.68 (0.21)\*\*** | -1.82 (2.90) | **19.44 (6.24)\*\*** | **39.36 (9.31)\*\*\*** | 26.10 (13.30) | **1.97 (0.54)\*\*\*** |
| Sex |  |  |  |  |  |  |
| Female | Reference | Reference | Reference | Reference | Reference | Reference |
| Male | 0.20 (0.17) | -6.22 (2.36)** | -4.23 (5.08) | -9.28 (7.63) | 20.73 (10.90) | -0.24 (0.44) |
| Type of resistance |  |  |  |  |  |  |
| DSTB | Reference | Reference | Reference | Reference | Reference | Reference |
| DRTB | 0.39 (0.20) | -2.36 (2.80) | 3.41 (6.04) | -4.52 (9.08) | 21.88 (12.96) | 0.79 (0.52) |
| **Covariance Parameter** |  |  |  |  |  |  |
| Individual Level | 0.33 (0.12) | 65.34 (23.27) | 409.99 (103.34) | 1224.86 (224.87) | 2392.22 (463.90) | 4.03 (0.76) |
| Residual | 1.50 (0.14) | 266.58 (25.64) | 940.21 (90.01) | 1349.07 (124.79) | 3067.18 (284.39) | 4.42 (0.42) |

\*p<0.05, \*\*p<0.01, \*\*\*p<0.001
<sup>a</sup>Defined by chest CT at the end of TB treatment
Abbreviations:
BMI – body mass index; DRTB – drug-resistant tuberculosis; DSTB – drug-susceptible tuberculosis; HbA1c – glycated hemoglobin; HDL – high-density lipoprotein; LDL – low-density lipoprotein; se – standard error; VAI – visceral adipose index
**Bold** indicated that the finding is statistically significant at $\alpha=0.05$

### Results from subgroup and sensitivity analyses

We did not observe significant differences in the relationship between BMI change and post-TB metabolic syndrome by either baseline BMI or diabetes status (p-values for multiplicative statistical interaction >0.05). Among individuals with BMI ≥18.5 kg/m^2^ at TB treatment initiation, the risk of having metabolic syndrome at the end of TB treatment was 13.6%, 21.6% at 6-months post-TB treatment, and 26.4% at 12-months post-TB treatment versus 0.0% at the end of TB treatment, 0% at 6-months post-TB treatment, and 10.7% at 12-months post-TB treatment among those with BMI <18.5 kg/m^2^ at TB treatment initiation (model did not converge) **(Table S3)**. The adjusted risk ratios for metabolic syndrome were slightly higher among those with diabetes compared to those without diabetes (aOR_diabetes_=1.79 vs. aOR_non-diabetes_=1.69, p-value=0.648) (**Table S4**).

In a model where we included self-reported prescription medications to manage metabolic diseases, the risk of having metabolic syndrome among those with ≥5% relative increase in BMI was 1.98 times (95%CI 1.02 – 3.85) the risk of those with <5% relative increase in BMI, adjusting for age, sex, drug-resistant types, and the clustering of metabolic syndrome data at the individual level (data not shown). In sensitivity analyses to account for bias due to covariate misspecification in multivariable models, the adjusted risk ratios comparing those with ≥5% relative increase in BMI to those with <5% relative increase in BMI ranged from 1.90 – 2.30 for metabolic syndrome **(Table S5)**.

## DISCUSSION

Among a well-characterized observational cohort of individuals successfully treated for pulmonary TB, we found that nearly half of the study participants had ≥5% relative increase in BMI during their course of TB treatment and that those with such an increase were more likely to have metabolic syndrome after TB treatment. Individuals who had ≥5% relative increase in BMI also had an HbA1c that was 0.37 points higher than those with <5% relative increase in BMI. We also found that individuals with the greatest risks of metabolic syndrome after successful TB treatment were those with a BMI ≥18.5 kg/m^2^ at TB treatment initiation or those with diabetes and a ≥5% relative increase in BMI during TB treatment. Because TB survivors have increased mortality rates even after successful TB treatment, and CVD accounts for the greatest proportion of post-TB deaths, [3, 4] improving CVD prevention among persons with TB should be a priority. Our study suggests that there was a high and increasing prevalence of metabolic syndrome in the post-TB population.

Weight loss is a characteristic symptom of TB disease, and undernutrition is a leading risk factor for TB. Weight gain during the first 2-3 months of TB treatment is typically associated with improved treatment outcomes. [5, 26–28] However, little is known about the long-term metabolic sequelae of rapid weight gain during TB treatment. In our cohort, individuals with ≥5% relative increase in BMI during TB treatment had double the risk of metabolic syndrome post-TB treatment. To our knowledge, this is the first study to estimate the relationship between BMI increase during TB treatment and post-TB cardiometabolic health outcomes. While rapid increases in BMI among adults are known to be associated with adverse CVD outcomes, [29] additional prospective studies and larger cohorts are warranted to confirm these findings, and also to determine whether TB disease amplifies the association between increases in BMI and cardiometabolic disease risk, as well as the mechanisms that underpin the relationship. These new studies will also need a non-TB control group to adequately evaluate how metabolic trajectories differ post-TB disease from the general population.

In our cohort, persons who had ≥5% relative increase in BMI had higher levels of post-TB HbA1c. Our findings also suggest that individuals with diabetes and ≥5% relative increase in BMI may have the highest risk of metabolic syndrome post-TB treatment, although this finding needs to be interpreted with caution due to the small sample size. Previous studies suggested that individuals with diabetes and persistent metabolic syndrome are more likely to develop micro-(e.g., retinopathy, nephropathy, neuropathy) or macro-vascular (e.g., cardiovascular diseases) complications[30]. If TB increases metabolic dysregulation among patients with TB and DM, this could increase the risk of diabetes complications, which are often irreversible and affect individuals’ disability adjusted life years and quality of life. Our findings reinforce existing recommendations for TB care to ensure linkages to diabetes and CVD care providers post-TB treatment. [31]

Nutrition is a key modifiable biological factor that influences both TB and cardiometabolic disease risk and is directly relevant to metabolic syndrome. In the context of TB treatment, nutritional interventions have gained increasing interest after a recent cluster-randomized controlled trial from India suggested that nutritional supplementation during TB treatment was associated with lower risk of TB mortality and reduced TB incidence among household contacts. [15, 32] However, it is critical to note that in India, undernutrition may be a key driver of TB incidence and poor TB treatment outcomes, which may not be the case for other settings, including Georgia. To date, post-TB interventions have focused on lung rehabilitation, cognitive behavioral therapy to address mental health, and hepatoprotective agents to prevent liver sequelae[33]. Currently, there are no guidelines or effective nutritional interventions to prevent poor cardiometabolic outcomes after TB treatment. Published studies suggested that nutritional interventions during TB treatment (e.g., micronutrient supplementations with zinc, retinol, or vitamin D; or general support including food baskets and food purchasing incentives) may improve treatment adherence and TB clinical outcomes, [34–36] but the long-term effects of these interventions remain unknown. Based on our findings, nutritional supplementation studies aimed at preventing CVD after TB treatment (e.g., adding medications that lower CVD risk like statins as adjuvant TB treatment) may be warranted. Such studies should evaluate whether nutritional interventions need to be targeted differently (e.g., setting a target priority, using different implementation strategies during TB treatment to prevent adverse TB outcomes and then modified to a different nutritional intervention after TB treatment to lower the risk of cardiometabolic diseases), especially in settings with different TB and CVD burdens.

Our study has several limitations. First, we only collected BMI data at two time points during TB treatment for the current study. Additional BMI or other biometric measures of nutritional status during and after TB treatment would allow better characterization of how changes in weight and nutritional status impact cardiometabolic disease risk. Second, we enrolled study participants at the end of TB treatment, limiting our ability to account for individuals’ cardiometabolic profiles pre-TB or at TB treatment initiation. Third, our follow-up period of 1-year post-TB treatment was relatively short. Longer follow-up periods after TB treatment completion are needed to better characterize the relationships between change in BMI during TB and cardiometabolic diseases. Fourth, we included TB patients with heterogeneous treatment regimens and durations that may introduce cohort effects. Fifth, cardiometabolic diseases are multi-factorial, and we did not have information on individuals’ physical activity, diet, family history, stress level, drugs/prescription records to indicate treatment adherence, and other risk factors that may influence prevalence/levels of post-TB metabolic syndrome or other cardiovascular/metabolic markers. We also did a single measurement of blood pressure at each study visit, which may cause spurious measurement of hypertension, leading to misclassifications of our primary outcome variable. Future studies aiming to estimate the prevalence/levels of post-TB metabolic syndrome or other cardiovascular/metabolic markers should consider including these important confounders and comparisons between different TB endotypes (e.g., newly diagnosed vs. recurrent TB, pulmonary vs. extrapulmonary TB, HIV co-infection status, structural lung disease, *Mtb* strains), which may help in characterizing hosts’ susceptibility, immunodeficiency, or pathologic inflammation profile. Lastly, we enrolled TB patients from one region in Georgia, and without non-TB individuals as a control group; the majority of whom were treated from the referral facility (i.e., NCTLD).

### Conclusion

In conclusion, we found that the risks of metabolic syndrome and elevated blood glucose levels post-TB treatment were increased among participants who had a BMI increase of at least 5% during TB treatment. Our results underscore the importance of following individuals after TB treatment and the need for clinicians to screen for cardiometabolic risks. In resource-limited settings with a high TB burden, integrating post-TB care within primary care clinics or other healthcare settings which have the capacity to do CVD risk assessment is critical, or alternatively, CVD screening could be integrated into TB care, especially among target priority groups (e.g., individuals with diabetes), and may reduce premature deaths after successful TB treatment.

## Data Availability

All data produced in the present study are available upon reasonable request to the authors

## DECLARATIONS AND ACKNOWLEDGEMENTS

### ETHICS APPROVAL

This study was submitted to, reviewed, and approved by the Institutional Review Boards (IRBs) at Emory University, Atlanta, USA, and the ethics committee at the National Center for Tuberculosis and Lung Diseases, Tbilisi, Georgia (FWA000020831). All participants provided written informed consent prior to study participation.

### AUTHOR CONTRIBUTIONS

MJM, MK, RRK, and ADS conceived the study design. TA, MG, and MK collected study data. ADS performed the analyses. ADS, HK, RRK, MJM, and MK interpreted the results. ADS wrote the first draft of the manuscript. MJM, RRK, and MK assisted with further drafting of the manuscript. All authors reviewed and approved the final version of the manuscript.

## ACKNOWLEGEMENTS

The authors would like to thank the study doctors (Manana Rekhviashvili, Khatuna Guchmazashvili, Marine Gachechiladze, Liana Tsivtsivadze, Tamar Natriashvili, Lia Ghambashidze) who helped the participant enrollment process at the National Center for Tuberculosis and Lung Diseases, Tbilisi, Georgia.

## SUPPLEMENTARY MATERIALS

**Figure S1.**
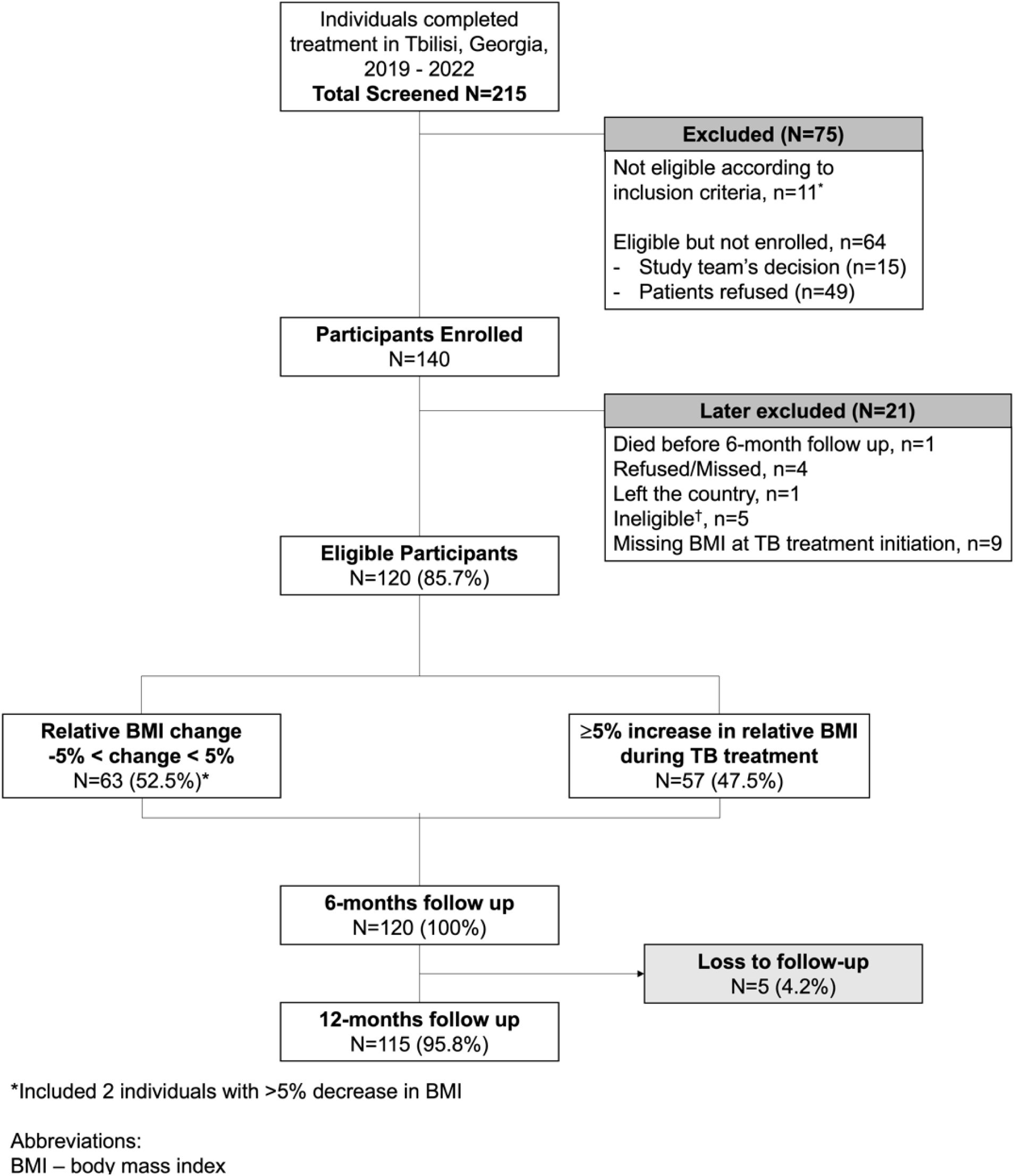
Study flow. **Alt text:** Flow diagram illustrating the sequential exclusion steps and study sample selection

**Figure S2.**
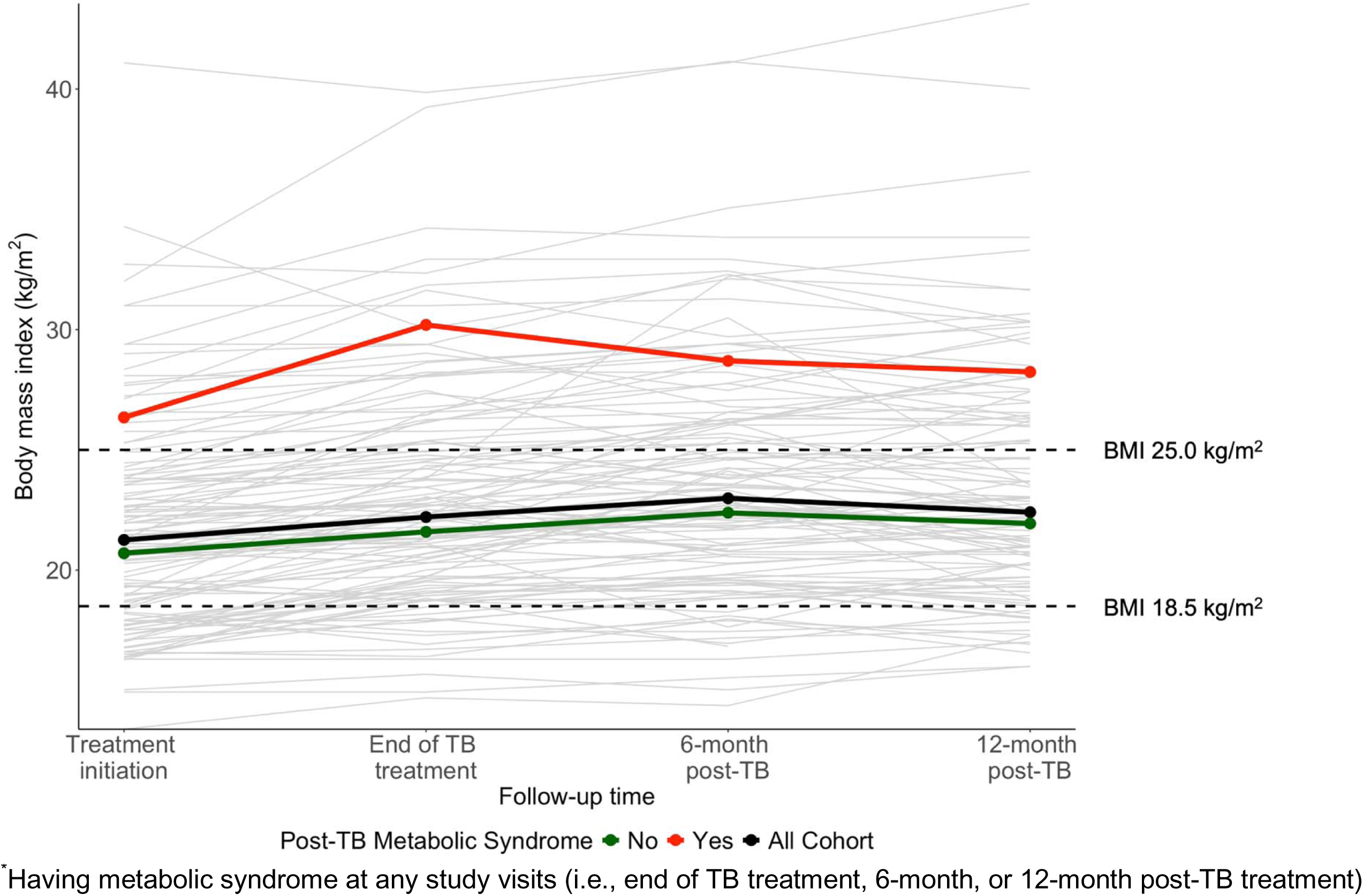
Body mass index trajectories from tuberculosis treatment initiation to 12-month post-treatment stratified by post-metabolic syndrome status* among individuals successfully treated for pulmonary tuberculosis, Georgia, 2020 - 2022 (N = 120) **Alt text:** A graph showing individual and group trajectories of BMI during and after TB treatment

**Table S1.** Prevalence of metabolic syndrome and disorders according to changes in body mass index among individuals who were successfully treated for pulmonary tuberculosis diseases in the country of Georgia, 2019 – 2022 (N=120)

| BMI Change | Prevalence (%) |  |  |
| --- | --- | --- | --- |
|  | T0 | T6 | T12 |
| **Metabolic Syndrome<sup>a</sup>** |  |  |  |
| All cohort | 10.0 | 15.8 | 22.6 |
| <5% increase | 7.9 | 7.9 | 16.1 |
| ≥5% increase | 12.3 | 24.6 | 30.2 |
| **Hypertension (130/80mmHg)** |  |  |  |
| All cohort | 34.2 | 34.5 | 37.4 |
| <5% increase | 30.2 | 33.3 | 43.6 |
| ≥5% increase | 38.6 | 35.7 | 30.2 |
| **Triglycerides ≥150mg/dL** |  |  |  |
| All cohort | 16.7 | 21.7 | 21.7 |
| <5% increase | 15.9 | 17.5 | 12.9 |
| ≥5% increase | 17.5 | 26.3 | 32.1 |
| **Lower HDL** |  |  |  |
| All cohort | 42.9 | 40.8 | 39.1 |
| <5% increase | 35.7 | 38.1 | 35.5 |
| ≥5% increase | 51.0 | 43.9 | 43.4 |
| **HbA1c ≥5.7%** |  |  |  |
| All cohort | 6.7 | 16.8 | 34.8 |
| <5% increase | 9.5 | 6.4 | 30.7 |
| ≥5% increase | 3.5 | 28.8 | 39.6 |
| **Abdominal Obesity<sup>b</sup>** |  |  |  |
| All cohort | 16.0 | 20.8 | 24.4 |
| <5% increase | 14.3 | 19.1 | 19.4 |
| ≥5% increase | 17.9 | 22.8 | 30.2 |
| <sup>a</sup> Having at least three out of five following health conditions (elevated blood pressure, elevated triglycerides, low high-density lipoprotein, elevated blood glucose, and abdominal obesity)<br><sup>b</sup> Waist circumference of ≥88cm in women and ≥102cm in men<br>Abbreviations:<br>BMI – body mass index<br>**Bold** indicated that the finding is statistically significant at $\alpha=0.05$ | | | |

**Table S2.** Means of post-treatment metabolic markers according to changes in body mass index among individuals who were successfully treated for pulmonary tuberculosis diseases in the country of Georgia, 2019 – 2022

| **BMI Change** | **Mean\*** |  |  |
| --- | --- | --- | --- |
|  | **T0** | **T6** | **T12** |
| **HbA1c** |  |  |  |
| All Cohort | 4.38 | 4.79 | 5.35 |
| <5% increase | 4.47 | 4.31 | 5.21 |
| ≥5% increase | 4.29 | 5.33 | 5.52 |
| **HDL** |  |  |  |
| All Cohort | 49.83 | 50.82 | 51.58 |
| <5% increase | 48.96 | 52.48 | 50.02 |
| ≥5% increase | 50.82 | 48.98 | 53.41 |
| **LDL** |  |  |  |
| All Cohort | 76.29 | 82.78 | 92.13 |
| <5% increase | 75.06 | 84.20 | 88.66 |
| ≥5% increase | 77.70 | 81.21 | 96.19 |
| **Total Cholesterol** |  |  |  |
| All Cohort | 192.90 | 195.43 | 196.15 |
| <5% increase | 186.60 | 197.02 | 191.22 |
| ≥5% increase | 199.86 | 193.66 | 201.92 |
| **Triglyceride** |  |  |  |
| All Cohort | 110.20 | 118.88 | 111.36 |
| <5% increase | 110.07 | 112.34 | 99.08 |
| ≥5% increase | 110.36 | 126.10 | 125.72 |
| **VAI** |  |  |  |
| All Cohort | 3.61 | 4.18 | 3.73 |
| <5% increase | 3.58 | 3.72 | 3.20 |
| ≥5% increase | 3.65 | 4.70 | 4.34 |
| *Arithmetic means were presented to be consistent with results from the general linear mixed models |  |  |  |
| Abbreviations:<br>BMI – body mass index; CI – confidence interval;<br>HbA1c – glycated hemoglobin; HDL – high-density lipoprotein; LDL – low-density lipoprotein; VAI – visceral adipose index |  |  |  |
| **Bold** indicated that the finding is statistically significant at $\alpha=0.05$ | | | |

**Table S3.**
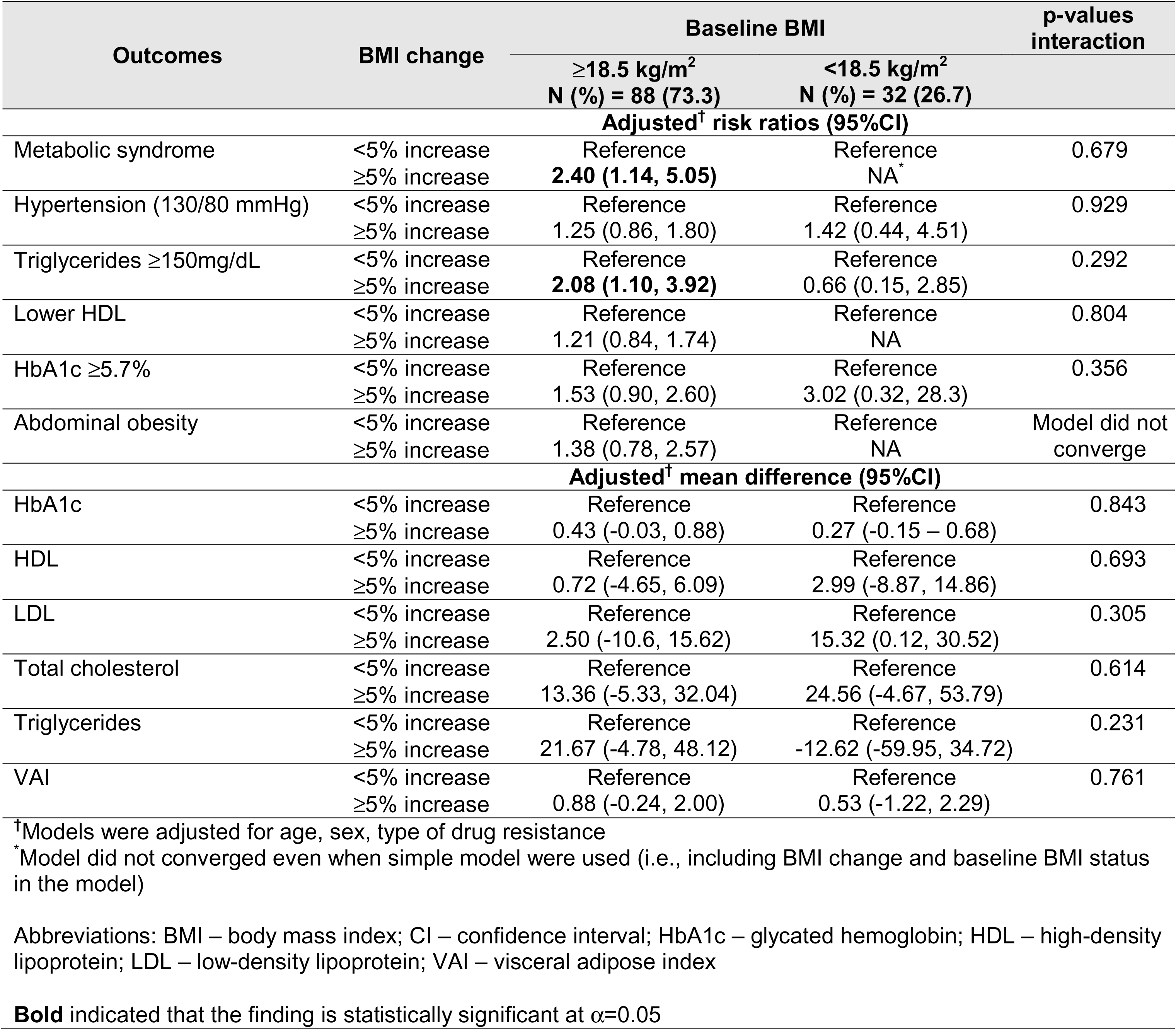
Statistical interaction between baseline BMI and BMI change during tuberculosis treatment on post-treatment metabolic markers among individuals who were successfully treated for pulmonary tuberculosis disease in the country of Georgia, 2019 – 2022

| Outcomes | BMI change | Baseline BMI |  | p-values<br>interaction |
| --- | --- | --- | --- | --- |
|  |  | ≥18.5 kg/m<sup>2</sup> | <18.5 kg/m<sup>2</sup> |  |
|  |  | N (%) = 88 (73.3) | N (%) = 32 (26.7) |  |
| Adjusted<sup>†</sup> risk ratios (95%CI) |  |  |  |  |
| Metabolic syndrome | <5% increase | Reference | Reference | 0.679 |
|  | ≥5% increase | **2.40 (1.14, 5.05)** | NA<sup>*</sup> |  |
| Hypertension (130/80 mmHg) | <5% increase | Reference | Reference | 0.929 |
|  | ≥5% increase | 1.25 (0.86, 1.80) | 1.42 (0.44, 4.51) |  |
| Triglycerides ≥150mg/dL | <5% increase | Reference | Reference | 0.292 |
|  | ≥5% increase | **2.08 (1.10, 3.92)** | 0.66 (0.15, 2.85) |  |
| Lower HDL | <5% increase | Reference | Reference | 0.804 |
|  | ≥5% increase | 1.21 (0.84, 1.74) | NA |  |
| HbA1c ≥5.7% | <5% increase | Reference | Reference | 0.356 |
|  | ≥5% increase | 1.53 (0.90, 2.60) | 3.02 (0.32, 28.3) |  |
| Abdominal obesity | <5% increase | Reference | Reference | Model did not<br>converge |
|  | ≥5% increase | 1.38 (0.78, 2.57) | NA |  |
| Adjusted<sup>†</sup> mean difference (95%CI) |  |  |  |  |
| HbA1c | <5% increase | Reference | Reference | 0.843 |
|  | ≥5% increase | 0.43 (-0.03, 0.88) | 0.27 (-0.15 – 0.68) |  |
| HDL | <5% increase | Reference | Reference | 0.693 |
|  | ≥5% increase | 0.72 (-4.65, 6.09) | 2.99 (-8.87, 14.86) |  |
| LDL | <5% increase | Reference | Reference | 0.305 |
|  | ≥5% increase | 2.50 (-10.6, 15.62) | 15.32 (0.12, 30.52) |  |
| Total cholesterol | <5% increase | Reference | Reference | 0.614 |
|  | ≥5% increase | 13.36 (-5.33, 32.04) | 24.56 (-4.67, 53.79) |  |
| Triglycerides | <5% increase | Reference | Reference | 0.231 |
|  | ≥5% increase | 21.67 (-4.78, 48.12) | -12.62 (-59.95, 34.72) |  |
| VAI | <5% increase | Reference | Reference | 0.761 |
|  | ≥5% increase | 0.88 (-0.24, 2.00) | 0.53 (-1.22, 2.29) |  |

<sup>†</sup>Models were adjusted for age, sex, type of drug resistance
\*Model did not converged even when simple model were used (i.e., including BMI change and baseline BMI status in the model)
Abbreviations: BMI – body mass index; CI – confidence interval; HbA1c – glycated hemoglobin; HDL – high-density lipoprotein; LDL – low-density lipoprotein; VAI – visceral adipose index
**Bold** indicated that the finding is statistically significant at $\alpha=0.05$

**Table S4.**
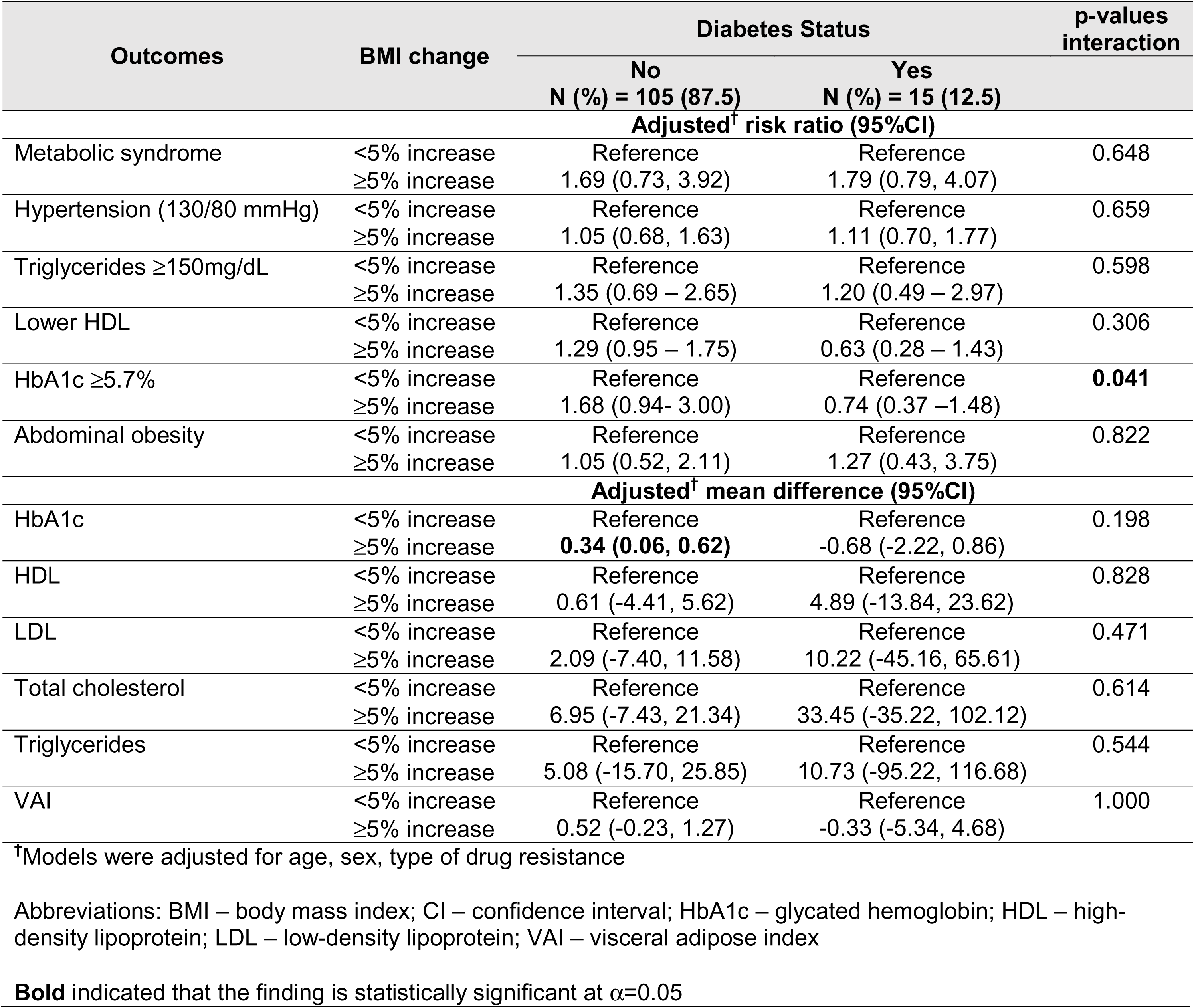
Statistical interaction between BMI change during tuberculosis treatment and diabetes status on post-treatment metabolic markers among individuals who were successfully treated for pulmonary tuberculosis disease in the country of Georgia, 2019 – 2022

**Table S5.** Statistical interaction between BMI change during tuberculosis treatment and diabetes status on post-treatment metabolic markers among individuals who were successfully treated for pulmonary tuberculosis disease in the country of Georgia, 2019 – 2022

| **Models** | **Covariates included** | **BMI Change** | **Metabolic Syndrome**<br>**RR (95%CI)** |
| --- | --- | --- | --- |
| Model 1 | BMI change | <5% increase<br>≥5% increase | Reference<br>**2.10 (1.12 – 3.94)** |
| Model 2 | BMI change, age, sex | <5% increase<br>≥5% increase | Reference<br>**2.30 (1.22 – 4.34)** |
| Model 3* | BMI change, age, sex, DST profile | <5% increase<br>≥5% increase | Reference<br>**2.07 (1.07 – 4.01)** |
| Model 4 | BMI change, age, sex, current smoker | <5% increase<br>≥5% increase | Reference<br>**2.07 (1.06 – 4.02)** |
| Model 5 | BMI change, age, sex, DST profile, current smoker | <5% increase<br>≥5% increase | Reference<br>1.90 (0.95 – 3.78) |
| *Model used in the manuscript<br>Abbreviations:<br>BMI – body mass index; CI – confidence interval; DST – drug sensitivity test; OR – odds ratio<br>**Bold** indicates that the finding is statistically significant at $\alpha=0.05$ |  |  |  |

